# *Mycobacterium tuberculosis* infection in pregnancy: a systematic review

**DOI:** 10.1101/2024.08.10.24311783

**Authors:** Alison J. Morton, Alexandra Roddy Mitchell, Richard E. Melville, Lisa Hui, Steven YC Tong, Sarah J. Dunstan, Justin T. Denholm

## Abstract

Pregnancy may be associated with risk of developing tuberculosis (TB) in those infected with *Mycobacterium tuberculosis* (*Mtb*). The perinatal period could provide opportunities for targeted screening and treatment. This study aims to synthesise published literature on *Mtb* infection in pregnancy, relating to prevalence, natural history, test performance, cascade of care, and treatment. We searched Ovid MEDLINE, Embase+Embase Classic, Web of Science, and Cochrane Central Register of Controlled Trials (CENTRAL) on October 3, 2023, and 47 studies met the inclusion criteria. The prevalence of *Mtb* infection was up to 57.0% in some populations, with rates increasing with maternal age and in women from high TB-incidence settings. Five studies quantified perinatal progression from *Mtb* infection to active TB disease, with two demonstrating increased risk compared to non-pregnant populations (IRR 1.3-1.4 during pregnancy and IRR 1.9-2 postpartum). Concordance between Tuberculin Skin Test (TST) and Interferon Gamma-Release Assay (IGRA) ranged from 49.4%-96.3%, with k-values of 0.19-0.56. High screening adherence was reported, with 62.0-100.0% completing antenatal TST and 81.0-100.0% having chest radiograph. Four studies of TB preventative treatment (TPT) did not find a significant association with serious adverse events. The antenatal period could provide opportunities for contextualised *Mtb* infection screening and treatment. As women with increased age and from high TB-incidence settings demonstrate higher prevalence and risk of disease, this cohort should be prioritised. TPT appears safe and feasible; however, further studies are needed to optimise algorithms, ensuring pregnant and postpartum women can make evidence-informed decisions for effective TB prevention.

## 1. Introduction

The impact of pregnancy on tuberculosis (TB) has been a subject of prolonged discourse since as early as P. Dutcher’s 1863 review in the Chicago Medical Journal [1]. Dutcher hypothesised that pregnancy “antagonises the development of disease” and may cause an increased risk of mortality [1]. Despite this early observation, 160 years later, we are still unsure of the true effect pregnancy has on TB, and pregnant women remain a neglected population in terms of evidence-based TB prevention [2-4].

TB is one of the top causes of death globally in women of reproductive age (15-45 years) and the leading non-obstetric cause of maternal mortality [5-8]. Active TB disease during pregnancy is associated with an increased risk for poor maternal and neonatal outcomes, including a twofold increase in maternal mortality [9, 10]. Therefore, diagnosing and treating active disease during the perinatal period offers clear benefits. Identifying and treating women with *Mtb* infection (immune sensitisation to *Mycobacterium tuberculosis*, without evidence of active TB disease) during the antenatal period could provide an opportunity to mitigate the risks of active TB disease in both the mother and neonate. The World Health Organization (WHO) currently recommends TB preventative treatment (TPT) to those at the highest risk of active TB disease [11]. This includes household contacts of those with active TB disease, people living with human immunodeficiency virus (HIV), and other high-risk groups whose immunity is compromised [11]. However, its value during pregnancy remains controversial. This uncertainty arises from the lack of data about the effectiveness of screening and treatment of *Mtb* infection in pregnant women [12, 13].

To further understand optimal management of *Mtb* infection in pregnancy, we conducted a systematic review addressing screening and treatment of *Mtb* infection, including yield and outcomes from testing and management.

## 2. Methods

### 2.1 Search Methodology

The protocol for this systematic review was registered with PROSPERO (CRD42023462868) on 26 September 2023. This study followed the Preferred Reporting Items for Systematic Reviews and Meta-analyses (PRISMA) reporting guidelines. Ovid MEDLINE, Embase+Embase Classic, Web of Science, and Cochrane Central Register of Controlled Trials (CENTRAL) were searched by AM from database inception to 3 October 2023. Full search terms are described in Appendix S1. Searches of electronic databases were supplemented by hand-searching the reference lists of included articles for further eligible studies. The search was restricted to studies written in English. There were no restrictions on the location of the study or publication date. Results from each database were imported into the reference management software EndNote™ 20 (Clarivate, Philadelphia, PA, USA). After removing duplicates, two reviewers (AM, ARM or RM) independently screened the titles and abstracts of retrieved studies. Two reviewers (AM and ARM) assessed full-text articles for eligibility. If consensus was not achieved, discrepancies were resolved by a third reviewer (JD). No patients were involved in the study process.

### 2.2 Study inclusion and exclusion criteria

The population of interest was pregnant/postnatal women (defined as the six months following birth) without known HIV infection. We excluded studies exclusively on pregnant women with HIV co-infection, given established indications for testing and treatment. For the purposes of this study, *Mtb* infection was defined as those who returned a positive tuberculin skin test (TST) or *Mtb*-specific interferon-γ release assay (IGRA) without evidence of active TB disease. We included studies reporting any aspect of *Mtb* infection in pregnancy, with broad co-primary outcomes of *Mtb* infection prevalence, natural history of progression to active TB disease, test performance, cascade of care, and *Mtb* infection treatment outcomes. Randomised control trials, cohort studies, case-control studies, cross-sectional studies, and descriptive studies were eligible for inclusion. Case reports, letters to the editor, commentaries, and conference abstracts were excluded. In cases where multiple publications used the same data, the most recent study was included, provided additional information was not included in earlier publications.

### 2.3 Data extraction

Two reviewers (AM and ARM) extracted data from eligible studies into a predefined Microsoft Excel data spreadsheet. Author(s), year of publication, country(ies), years of study recruitment, study design, sample size, included outcomes, assessment methods for outcomes, and outcomes were recorded. Any discrepancies were resolved following discussion with a third reviewer (JD).

### 2.4 Risk of bias (quality assessment)

Two reviewers (AM and ARM) independently assessed the included studies’ risk of bias. We used an eight-point checklist adapted from the Newcastle-Ottawa instrument (Appendix S2). One point was assigned for each checklist item, and the overall study quality score was calculated based on the sum of these points. We considered scores of 0 to 2 to be poor quality, 3 to 5 to be fair quality, and 6 to 8 to be good quality.

## 3. Results

### 3.1 Search results

The initial search produced 9,970 results after the removal of duplicates. After title and abstracts were screened, 117 full-text articles were reviewed. Six articles could not be retrieved. Two additional studies were identified following a manual search of the reference lists, resulting in a total of 47 articles included. This includes 25 cross-sectional studies, 17 cohort studies, two case-control studies, two open-label trials, and one qualitative study. Of these, 31 reported prevalence of *Mtb* infection in pregnancy, 20 investigated the cascade of care, 13 examined test performance of TST and IGRA in the perinatal period, eight addressed the natural history of progression to TB disease in the perinatal period, and four reported on treatment safety in the perinatal period. Some studies investigated more than one of these subjects. 31 studies reported from low TB incidence settings (<100 cases of TB per 100,000 individuals), while 16 reported from high TB incidence settings (≥100 cases of TB per 100,000 individuals). Characteristics of the included articles are summarised in Appendix S3.

**Figure 1.**
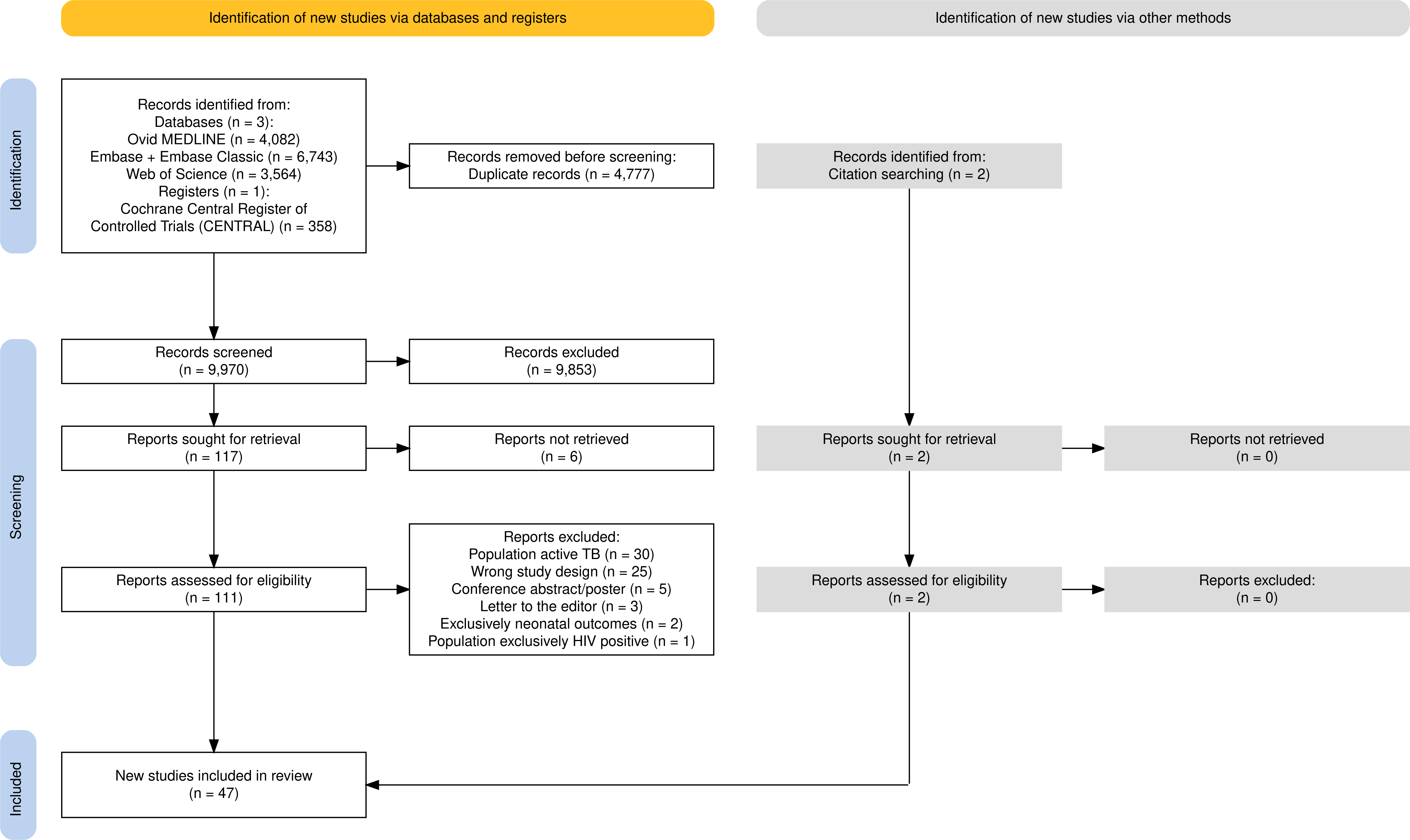
**PRISMA Flow Diagram**

### 3.2 Co-primary outcomes

#### 3.2.1 Prevalence

Thirty-one studies reported the prevalence of *Mtb* infection among pregnant women, using either TST or IGRA (Table 1). The reported prevalence rates varied substantially, ranging from 0.0% to 57.0% [14-44]. Of the included studies, twenty-one were conducted in low TB incidence settings (prevalence ranging from 0.0-50.4%), while ten were from high TB incidence settings (prevalence ranging from 9.6-57.0%).

**Table 1.**
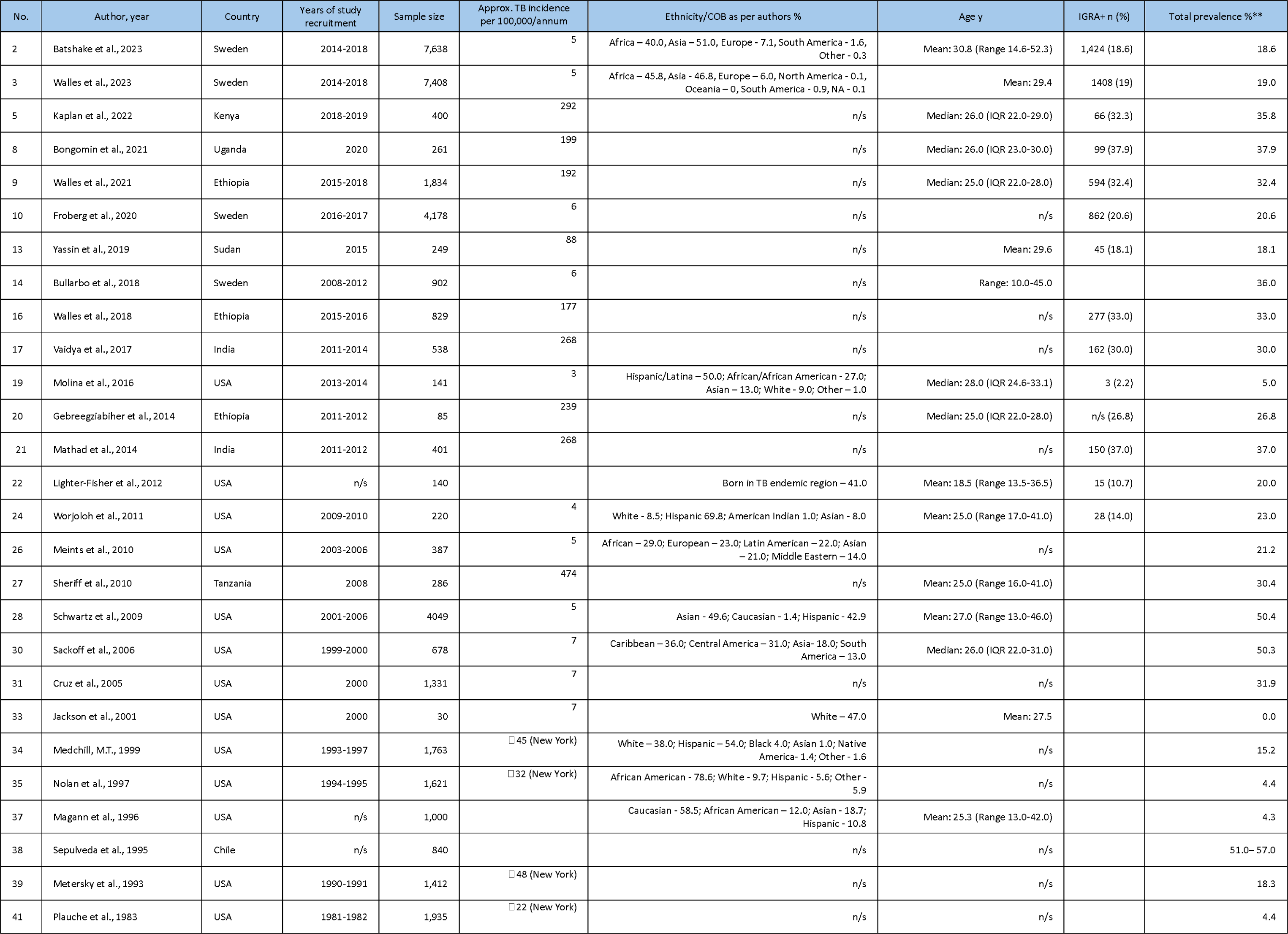
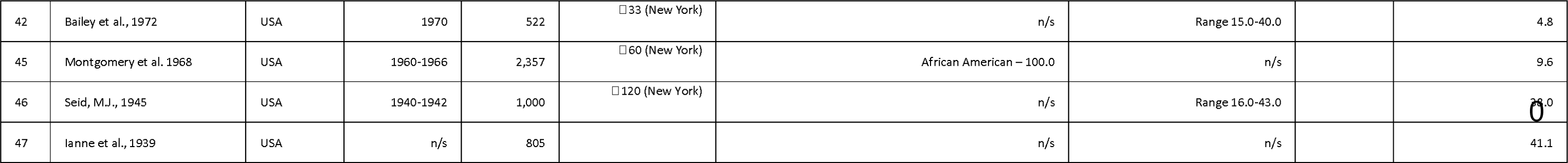
Prevalence of Mtb Infection During Pregnancy Measured by TST or IGRA. IQR = Interquartile range; HIV+ = Human Immunodeficiency Virus seropositive; BCG= Bacille Calmette-Guérin; TST = Tuberculin skin test; IGRA = Interferon-Gamma Release Assay; n/s= not stated *Manufacturer cut-offs were used for all studies **Positive by either IGRA or TST

Six studies found a positive association between *Mtb* infection and increasing maternal age. In one study from Uganda, the prevalence of *Mtb* infection was 37.9%, based on a positive QuantiFERON-TB Gold Plus (QFT-Plus; a commercial IGRA) result [17]. On multivariate analysis, pregnant women between 30-39 years of age were four times more likely to have *Mtb* infection than those below 20 years (adjusted OR, 3.96 95% CI 1.23–12.73; P = .021) [17]. This finding is congruent with a similar study from Ethiopia. With an overall reported prevalence of *Mtb* infection at 32.4%, 20.0% of participants seeking antenatal care had acquired *Mtb* infection at the age of 18 years [18]. This proportion increased to 46.0% among those aged over 26 years, implying continued TB transmission in the communities where these women live [18].

Ten studies found a positive association between *Mtb* infection and ethnicity or country of birth. In one cross-sectional study from the United States, 31.3% of Asian American women, 23.9% of South American women, 9.3% of African American women, and 4.1% of Caucasian women had a positive TST [35]. In this study, the calculated relative risk of *Mtb* infection in relation to ethnicity revealed that women of South American origin had a risk ratio of 5.9 (95% CI 3.9 to 8.8), while Asian-American women had a risk ratio of 7.6 (95% CI 3.4 to 17.5), both compared to Caucasian women [35]. In a retrospective cohort study in Sweden, the prevalence of perinatal *Mtb* infection was 18.6% [45]. Among women originating from Africa, 27.4% were positive on IGRA, compared to 12.9% from Asia and 10.7% from Europe [45]. Compared to all other world regions, *Mtb* infection was significantly more common among women of African origin (27.4% vs 12.7%; P < .001) [45].

#### 3.2.2 Natural history

Eight studies examined whether pregnancy and the postnatal period constitute an independent risk factor for the development of active TB disease. Earlier retrospective studies from the Dominican Republic in 1998, Malawi in 2004 and the United States in 1956 suggested no increased risk of TB activation during pregnancy and the postnatal period [46-48]. Since 2012, five studies have quantified the risk of progression to TB disease in the perinatal period, with two studies demonstrating increased incidence risk ratios when compared to non-pregnant populations (IRR 1.3-1.4 during pregnancy and IRR 1.9-2 postpartum) [49, 50].

In a retrospective cohort study by Zenner et al., utilising the General Practitioner Research Database, the incidence of TB diagnosis was notably elevated postpartum [49]. The study included 192,801 women with 264,136 pregnancies in the UK between 1996 and 2008. The crude incidence rate of active TB in the general UK population was 10.1/100,000 person-years (95% CI 8.7–11.8) [49]. During pregnancy, the rate was 12.8/100,000 person-years (95% CI 8–19.4), and during the 180-day postpartum period was 19.2/100,000 person-years (95% CI 12–29) [49]. After adjusting for age, socioeconomic status, region of residence, and bacille Calmette-Guérin (BCG) vaccination, the incidence of active TB disease was significantly higher during the 180-day postpartum period (IRR, 1.95 [95% CI 1.24–3.07]), but not during pregnancy (IRR, 1.29 [95% CI 0.82-2.03]) [49]. The authors suggested that the timing of postpartum diagnosis may reflect antepartum TB disease onset [49].

In a retrospective cohort study in Sweden covering registered deliveries from 2005 to 2013, an increased risk of active TB was observed during pregnancy and the postpartum period [50]. The cohort included 649,342 women with a total of 951,530 deliveries. During the study, 553 women were diagnosed with active TB, with 85 cases during pregnancy, 79 during postpartum, and 389 when not pregnant or postpartum [50]. The overall risk of active TB increased during pregnancy (IRR 1.4, 95% CI 1.1–1.7) and postpartum (IRR 1.9, 95% CI 1.5–2.5) compared to when not pregnant or postpartum [50].

Since 2016, three further studies have demonstrated an increased risk of progression to TB disease in the perinatal period. In another retrospective study from Sweden, 14 of 1,424 enrolled women developed TB disease during the perinatal period, corresponding to an incidence rate of pregnancy-related Mtb infection progression to TB disease of 7.8 (95% CI, 4.6–13.2) cases per 1,000 person-years [45]. Within a prospective study in India, nine pregnant women progressed to active TB disease [51]. The overall TB incidence was 28 per 1,000 person-years (95%, CI 13 – 53), which is 24.6 times higher than the TB incidence of 1.14 per 1,000 person-years reported from the general population in the same state [51]. A study from Mongolia illustrated that pregnant women had a 1.3-fold higher risk of developing TB than the general population (IRR 1.31 [95%CI, 1.081.59]) [52].

#### 3.2.3. TST and IGRA performance for indicating Mtb infection

Thirteen studies examined the impact of immune changes during the perinatal period on the TST and IGRA diagnostic performance.

In eight comparative studies between TST and IGRA, concordance ranged from 49.4 to 96.3%, with kappa values of 0.19 to 0.56 [16, 24, 26, 27, 51, 53, 54]. Discordance, most commonly TST positive/IGRA negative, was observed in high TB-incidence settings, with authors suggesting that this was likely linked to BCG-vaccinated populations and increased exposure to non-tuberculous mycobacteria (NTM) [24, 27, 28]. Two studies reported IGRA positive/TST negative discordance in pregnant women. The authors concluded that this most likely reflected ‘false-negative’ TST results, as IGRA results more closely mirrored the general population of each respective setting [26]. Reasons given by the authors for ‘false-negative’ TST results within these studies included anergy, recent Mtb infection and incorrect TST interpretation [16].

One cohort study in India revealed a pregnancy-related impact on interferon-gamma response to M. tuberculosis stimulation. IGRA positivity decreased at delivery (100% to 77%, p<0.01), followed by an increase between delivery and six months postpartum (77% to 89%, p<0.05) [51]. The authors propose that these fluctuations signify immune changes during pregnancy [51]. Thus, they conclude that the immune alterations in pregnancy and the postpartum period impact the performance of IGRA, potentially leading to false-negative test results in late pregnancy [51].

#### 3.2.4 Cascade of care

Twenty studies focused on the cascade of care, from antenatal TST/IGRA screening to TPT completion (Table 2). Reported antenatal TST adherence ranged from 62-100%, and chest radiography 81-100%. TPT completion rates varied, with a higher likelihood of women successfully completing treatment during the antenatal period [19, 32, 55, 56]. Those studies with high treatment completion rates stated possible reasons may include intensive nurse case management [19, 55, 57]. In a Swedish study examining the cascade of care for pregnant women with a positive IGRA, 439 women from high TB-incidence countries were included [57]. From antenatal screening, 177 women were recommended TPT [57]. Of these, 137 women received TPT, and 112 completed the course [57]. Only three out of 137 treated women were lost to follow-up (2%) [57]. The study found no association between treatment completion and treatment regimens [57]. The authors conclude that antenatal screening of pregnant women from high TB-incidence countries appears to be an effective and valuable strategy for identifying cases of Mtb infection [57].

**Table 2.** Perinatal Screening for Mtb Infection Cascade of Care. TST = Tuberculin skin test; IGRA = Interferon-Gamma Release Assay; CXR = Chest radiograph; 9H = 9 months daily isoniazid; 6H = 6 months of daily isoniazid; 4R = 4 months of daily rifampicin; 3HP = 3-month once-weekly isoniazid-rifapentine; 3HR = 3 months of daily isoniazid plus rifampin; 12H = 12 months of daily isoniazid; H = isoniazid

| No. | Author, year | Country/ies | Years of study recruitment | Sample size n | TST placed n | TST read n (%) | IGRA conducted n | Adherence to CXR n (%) | Participants commenced on treatment n (%) | Treatment regimen/s | Completion of treatment n (%) |
| --- | --- | --- | --- | --- | --- | --- | --- | --- | --- | --- | --- |
| 1 | Arvidsson et al., 2023 | Sweden | 2013-2018 | 439 |  |  | 439 | 421 (95.9) | 137 (31.9) | 9H, 6H, 4R | 112 (81.8) |
| 6 | Mathad et al., 2022 | Haiti, Kenya, Malawi, Thailand, and Zimbabwe | 2017-2018 | 50 |  |  |  |  | 50 (100.0) | 3HP | 50 (100.0) |
| 10 | Froberg et al., 2020 | Sweden | 2016-2017 | 4,178 |  |  | 4,178 | n/s | 255 (34.3) | 4R, 9H, 3HR, 3HP | 234 (91.8) |
| 14 | Bullarbo et al., 2018 | Sweden | 2008-2012 | 902 | 902 | 902 (100.0) |  | 306 (93.0) | 0 (0.0) |  |  |
| 18 | Molina et al., 2016 | USA | 2013-2014 | 143 | 137 | 85 (62.0) | 134 | 9 (100.0) |  |  |  |
| 20 | Mathad et al., 2014 | India | 2011-2012 | 401 | 401 | 352 (87.0) | 401 |  |  |  |  |
| 23 | Worjolah et al., 2011 | USA | 2009-2010 | 220 | 220 | 199 (95) |  |  |  |  |  |
| 26 | Sheriff et al., 2010 | Tanzania | 2008 | 286 | 286 | 286 (100.0) |  | 286 (100.0) |  |  |  |
| 27 | Schwartz et al., 2009 | USA | 2001-2006 | 4049 | 4,049 | 3,847 (95.0) |  | 1,841 (95.0) |  |  |  |
| 28 | Kwara et al., 2008 | USA | 2003 | 845 |  |  |  | 845 (100.0) | 690 (81.7) | 9H | 426 (61.7) |
| 29 | Sackoff et al., 2006 | USA | 1999-2000 | 730 | 521 | 521 (100.0) |  | n/s | 291 (100.0) | 6H | 27 (9.3) |
| 30 | Cruz et al., 2005 | USA | 2000 | 1,331 | n/s | 1,195 (90.0) |  | n/s | n/s | 6H | 71 (42.0) |
| 31 | Medchill, M.T., 1999 | USA | 1993-1997 | 1,763 | 1,634 | 1,497 (92.0) |  | 211 (93.0) |  |  |  |
| 38 | Metersky et al., 1993 | USA | 1990-1991 | 1,412 | 1405 | 1405 (100) |  | 254 (98.0) | n/s | 6H | 2 (5.0) |
| 39 | Franks et al., 1989 | USA | 1981-1982 | 3,681 |  |  |  |  | 3,681 (100.0) | 12H | 589 (16.0) |
| 40 | Plauche et al., 1983 | USA | 1981-1982 | 1,935 | 1,935 | 1,142 (59.0) |  | 85 (100.0) |  |  |  |
| 42 | Bailey et al., 1972 | USA | 1970 | 522 | 522 | 522 (100.0) |  | 25 (100.0) | 13 (52.0) | H |  |
| 43 | Montgomery et al. 1968 | USA | 1960-1966 | 2,643 | 2,643 | 2,357 (89.2) |  | 142 (100.0) |  |  |  |
| 45 | Seid, M.J., 1945 | USA | 1940-1942 | 1,000 | 1,000 | 988 (98.8) |  | 841 (100.0) |  |  |  |
| 46 | lanne et al., 1939 | USA | n/s | 805 | 805 | 691 (85.8) |  | 252 (87.0) |  |  |  |

One study examined predictors of TPT completion in the United States. Treatment initiation during the postpartum period was negatively associated with treatment completion [56]. Only 52% of pregnant women referred for postpartum therapy returned to initiate it, and only a third of those who started therapy postpartum completed it [56]. The authors underscore the potential value of the antenatal period as a window for optimising treatment completion [56].

One qualitative study was included, exploring the lived experience of pregnant women with a positive IGRA. Results from this study show that a positive IGRA initially caused women significant distress, as it was misunderstood as being active TB [58]. However, participants subsequently understood their condition well and were highly motivated to take TPT [58].

#### 3.2.4 Preventative Treatment

Four studies examined the safety of TPT during the perinatal period. In an open-label medication trial, pregnancy outcomes were evaluated for 125 women inadvertently exposed to three months of weekly isoniazid and rifapentine or nine months of daily isoniazid [59]. The study revealed no treatment-related maternal or neonatal serious adverse events (SAEs) [59]. This provided reassurance to clinicians and patients that these regimens are the safest option compared to other regimes where there was no data available for treatment during pregnancy [59]. A retrospective cohort study from Sweden analysed treatment outcomes of 439 pregnant women taking both isoniazid and rifampicin-based regimens [57]. Again, no maternal or neonatal SAEs were reported [57]. However, the primary reasons for treatment discontinuation were liver-related side effects (5.1%) and other treatment-related side effects (5.1%) [57]. The study lacked further information on the severity or grading of these side effects.

A retrospective cohort study assessed 3,681 pregnant and postpartum women participating in an isoniazid (INH) TPT program in the United States [60]. Five cases of INH hepatitis (0.14%) were identified, and two women died (0.05%) postpartum in those who received INH [60]. Ten cases of INH hepatitis (0.25%) and one maternal death (0.03%) were also reported in the non-pregnant comparison group [60]. Analysis raised the possibility of associations between pregnancy and the post-partum period with INH hepatitis (risk ratio of 2.5, 95% CI 0.8–8.2) and fatal hepatotoxicity (rate ratio 4.95% CI 0.2–258) [60]. However, the authors conclude that the increased risk was not statistically significant [60].

A non-randomised open-label pharmacokinetic and safety study of three months of weekly isoniazid and rifapentine (3HP) concluded that this regimen does not require dose adjustment in pregnancy [55]. All 50 women achieved exposures of rifapentine and isoniazid associated with successful tuberculosis prevention [55]. None of the reported SAEs within the trial were deemed to be related to treatment, and the data supports proceeding with larger safety-focused studies of 3HP in pregnancy [55].

### 3.3 Bias

This systematic review identified 47 studies. On quality assessment, 31 studies (66.0%) were of good quality, 14 (29.8%) were of fair quality, and 2 (4.2%) were of poor quality (Appendix S3).

## 4. Discussion

### 4.1 Main findings

This systematic review examined screening and treatment of Mtb infection among pregnant women. We found that the reported prevalence of Mtb infection varied widely between cohorts, with the highest risk observed in settings with a high background risk of tuberculosis, among individuals originating from regions with high TB incidence, and with increasing maternal age. Pregnancy and the post-partum period were associated with an increased risk of progression to TB disease in five studies, while three studies found no association. Pregnancy does not appear to affect concordance between the Tuberculin Skin Test (TST) and Interferon Gamma Release Assay (IGRA). Screening programs in pregnant populations revealed good completion of initial antenatal screening and chest radiograph. While treatment completion rates varied, women were more likely to successfully complete treatment during the antenatal period. TPT during the perinatal period had no associated SAEs in three studies, while one study reported a possible association with isoniazid hepatitis and fatal hepatotoxicity.

High proportions of Mtb infection were found in studies performed in both high and low-incidence countries, reflecting the algorithms used to select women for testing. This is reflective of likely real-world application. While some high-incidence settings may benefit most from universal screening and treatment programs, low-incidence settings are likely to develop algorithms for selection of higher-risk individuals (particularly based on country of birth and age). Data synthesised here can support the development and evaluation of evidence-based, contextualised screening strategies to maximise benefit.

This manuscript updates a previous 2016 review of Mtb infection in pregnancy [61]. Our study includes 25 additional studies, including six published before 1980 and nineteen since April 30, 2014. Of note, four studies since 2016 have added further epidemiological evidence to support the hypothesis that the perinatal period constitutes an independent risk factor for the progression to TB disease [45, 50-52]. Furthermore, we included three studies on Mtb infection treatment during the perinatal period [55, 57, 59]. Through a review of the available open-label trials and retrospective cohort studies, evidence supports the safety of several agents for TPT in pregnancy [55, 57, 59]. The conclusion of these clinical trials is at odds with the earlier, widely cited retrospective cohort study of Franks et al. [60]. Within this study, there was a non-significant association between pregnancy and the post-partum period with INH hepatitis and fatal hepatotoxicity [60]. It is worth noting that the adverse events occurred under normal programmatic conditions; the pregnant and non-pregnant groups were unmatched, followed a decade apart, and assessment of risk factors for INH-induced hepatitis (e.g. alcohol consumption, baseline elevation of transaminases and viral hepatitis screen) was inconsistent [60]. Including these newer studies in this updated systematic review provides further evidence supporting the safety of TPT in pregnancy. However, additional safety studies are warranted to substantiate the existing evidence. Prioritising the inclusion of pregnant and breastfeeding women in TB medication trials is crucial to quantify the safety and efficacy of TPT for both mothers and neonates.

The results of this review highlight the current missed opportunity the antenatal period provides for Mtb infection screening and treatment. Beyond the direct advantages for women and neonates to potentially reduce the risk of active TB disease, targeting Mtb infection is a crucial component in the broader strategy of TB control and elimination [62, 63]. Evidence suggests that TPT in non-pregnant at-risk populations reduces the risk of developing active TB disease; however, further studies should be prioritised to quantify the benefits of TPT within pregnant populations [64].

Our review identified only one qualitative study exploring the lived experience of pregnant women screened and treated for Mtb infection [58]. Further mixed methods analyses would be instrumental in identifying attitudes, behaviours, challenges, and opportunities, offering valuable insights for shaping effective interventions to enhance the care of pregnant women with Mtb infection.

### 4.2 Strengths

This systematic review was based on a pre-prepared protocol, and PRISMA guidelines were followed. The selected articles were retrieved via a comprehensive electronic search strategy from database inception to October 3, 2023. Additionally, no restriction was placed upon the study setting, allowing for a thorough overview of Mtb infection in pregnancy across low and high TB-incidence settings. Study selection, data extraction and risk-of-bias assessment performed independently by two reviewers reduced individual bias.

### 4.3 Limitations

Only published original studies were included in this systematic review, and the exclusion of grey literature may have resulted in the omission of relevant data. The studies included were of high methodological heterogeneity and covered multiple aspects of Mtb infection; therefore, no meta-analysis for each outcome could be performed. Finally, a broader limitation pertains to the challenge of generalising this study’s conclusions across different countries, given the inherent disparities in resources and healthcare infrastructure.

## 5. Conclusion

This study found that the antenatal period could provide an opportunity for targeted screening and treatment for pregnant women with Mtb infection. While in high TB incidence settings, universal screening may be warranted, these findings may also assist in risk-stratified targeting of screening in low-incidence settings. As women with increased maternal age and those from high TB-incidence settings demonstrate the highest prevalence and risk of disease, this cohort should be prioritised. Pregnancy does not appear to affect concordance between the TST or IGRA, and screening programs in pregnant populations revealed good antenatal adherence. From the limited number of published studies, antenatal TPT appears safe and feasible; however, further studies are needed to optimise benefits, ensuring pregnant and postpartum women can make evidence-informed decisions for effective TB prevention.

## Supporting information

S1 Search Strategy

S2 Modified NOS

S3 Characteristics of the Included Studies

S4 Table 1 Prevalence

## Data Availability

All data produced in the present work are contained in the manuscript

## Acknowledgements

The authors have no additional contributors to acknowledge.

## Notes

### Competing Interest Statement

The authors have declared no competing interest.

### Funding Statement

No funding was received for this study

