## Supplementary material for "*Mycobacterium tuberculosis* infection in pregnancy: a systematic review": S1 Search Strategy

**Appendix S1. Search Strategy**

**Ovid MEDLINE**

| **Search Number**  Results | Search Terms |
| --- | --- |
| **1**  1406004 | pregnancy/ OR puerperium/ OR parturition/ OR postpartum period/ OR antepartum period/ OR obstetrics/ |
| **2**  1142371 | pregnan*.mp. [mp=title, abstract, original title, name of substance word, subject heading word, floating sub-heading word, keyword heading word, organism supplementary concept word, protocol supplementary concept word, rare disease supplementary concept word, unique identifier, synonyms] |
| **3**  26832 | puerper*.mp. [mp=title, abstract, original title, name of substance word, subject heading word, floating sub-heading word, keyword heading word, organism supplementary concept word, protocol supplementary concept word, rare disease supplementary concept word, unique identifier, synonyms] |
| **4**  37497 | parturi*.mp. [mp=title, abstract, original title, name of substance word, subject heading word, floating sub-heading word, keyword heading word, organism supplementary concept word, protocol supplementary concept word, rare disease supplementary concept word, unique identifier, synonyms] |
| **5**  87502 | postpartum.mp. [mp=title, abstract, original title, name of substance word, subject heading word, floating sub-heading word, keyword heading word, organism supplementary concept word, protocol supplementary concept word, rare disease supplementary concept word, unique identifier, synonyms] |
| **6**  6801 | antepartum.mp. [mp=title, abstract, original title, name of substance word, subject heading word, floating sub-heading word, keyword heading word, organism supplementary concept word, protocol supplementary concept word, rare disease supplementary concept word, unique identifier, synonyms] |
| **7**  209183 | obstetric*.mp. [mp=title, abstract, original title, name of substance word, subject heading word, floating sub-heading word, keyword heading word, organism supplementary concept word, protocol supplementary concept word, rare disease supplementary concept word, unique identifier, synonyms] |
| **8**  298331 | mycobacterium tuberculosis/ OR tuberculosis/ OR latent tuberculosis/ OR tuberculosis infection/ |
| **9**  75815 | mycobacterium tuberculosis.mp. [mp=title, abstract, original title, name of substance word, subject heading word, floating sub-heading word, keyword heading word, organism supplementary concept word, protocol supplementary concept word, rare disease supplementary concept word, unique identifier, synonyms] |
| **10**  279433 | tuberculosis.mp. [mp=title, abstract, original title, name of substance word, subject heading word, floating sub-heading word, keyword heading word, organism supplementary concept word, protocol supplementary concept word, rare disease supplementary concept word, unique identifier, synonyms] |
| **11**  6430 | latent tuberculosis.mp. [mp=title, abstract, original title, name of substance word, subject heading word, floating sub-heading word, keyword heading word, organism supplementary concept word, protocol supplementary concept word, rare disease supplementary concept word, unique identifier, synonyms] |
| **12**  10060 | tuberculosis infection.mp. [mp=title, abstract, original title, name of substance word, subject heading word, floating sub-heading word, keyword heading word, organism supplementary concept word, protocol supplementary concept word, rare disease supplementary concept word, unique identifier, synonyms] |
| **13**  1475334 | #1 OR #2 OR #3 OR #4 OR #5 OR #6 OR #7 |
| **14**  298331 | #8 OR #9 OR #10 OR #11 OR #12 |
| **15**  5506 | #13 AND #14 |
| **16**  4082 | limit 15 to English language |
| Total | 4082 |

**Embase + Embase Classic**

| **Search Number**  Results | Search Terms |
| --- | --- |
| **1**  1604687 | pregnancy/ OR puerperium/ OR parturition/ OR postpartum period/ OR antepartum period/ OR obstetrics/ |
| **2**  1271367 | pregnan*.mp. [mp=title, abstract, original title, name of substance word, subject heading word, floating sub-heading word, keyword heading word, organism supplementary concept word, protocol supplementary concept word, rare disease supplementary concept word, unique identifier, synonyms] |
| **3**  76685 | puerper*.mp. [mp=title, abstract, original title, name of substance word, subject heading word, floating sub-heading word, keyword heading word, organism supplementary concept word, protocol supplementary concept word, rare disease supplementary concept word, unique identifier, synonyms] |
| **4**  33602 | parturi*mp [mp=title, abstract, original title, name of substance word, subject heading word, floating sub-heading word, keyword heading word, organism supplementary concept word, protocol supplementary concept word, rare disease supplementary concept word, unique identifier, synonyms] |
| **5**  102899 | postpartum.mp. [mp=title, abstract, original title, name of substance word, subject heading word, floating sub-heading word, keyword heading word, organism supplementary concept word, protocol supplementary concept word, rare disease supplementary concept word, unique identifier, synonyms] |
| **6**  11281 | antepartum.mp. [mp=title, abstract, original title, name of substance word, subject heading word, floating sub-heading word, keyword heading word, organism supplementary concept word, protocol supplementary concept word, rare disease supplementary concept word, unique identifier, synonyms] |
| **7**  236947 | obstetric*.mp [mp=title, abstract, original title, name of substance word, subject heading word, floating sub-heading word, keyword heading word, organism supplementary concept word, protocol supplementary concept word, rare disease supplementary concept word, unique identifier, synonyms] |
| **8**  374583 | mycobacterium tuberculosis/ OR tuberculosis/ OR latent tuberculosis/ OR tuberculosis infection/ |
| **9**  106953 | mycobacterium tuberculosis.mp. [mp=title, abstract, original title, name of substance word, subject heading word, floating sub-heading word, keyword heading word, organism supplementary concept word, protocol supplementary concept word, rare disease supplementary concept word, unique identifier, synonyms] |
| **10**  356635 | tuberculosis.mp. [mp=title, abstract, original title, name of substance word, subject heading word, floating sub-heading word, keyword heading word, organism supplementary concept word, protocol supplementary concept word, rare disease supplementary concept word, unique identifier, synonyms] |
| **11**  10221 | latent tuberculosis.mp. [mp=title, abstract, original title, name of substance word, subject heading word, floating sub-heading word, keyword heading word, organism supplementary concept word, protocol supplementary concept word, rare disease supplementary concept word, unique identifier, synonyms] |
| **12**  13017 | tuberculosis infection.mp. [mp=title, abstract, original title, name of substance word, subject heading word, floating sub-heading word, keyword heading word, organism supplementary concept word, protocol supplementary concept word, rare disease supplementary concept word, unique identifier, synonyms] |
| **13**  1752756 | #1 OR #2 OR #3 OR #4 OR #5 OR #6 OR #7 |
| **14**  374583 | #8 OR #9 OR #10 OR #11 OR #12 |
| **15**  8091 | #13 AND #14 |
| **16**  6743 | limit 15 to English language |
| Total | 6743 |

**Web of Science**

| **Search Number**  Results | Search Terms |
| --- | --- |
| **1**  967,790 | (((((ALL=(pregnan*)) OR ALL=(puerper*)) OR ALL=(parturi*)) OR ALL=(postpartum)) OR ALL=(antepartum)) OR ALL=(obstetric*) |
| **2**  243,883 | (((ALL=(mycobacterium tuberculosis)) OR ALL=(tuberculosis)) OR ALL=(latent tuberculosis)) OR ALL=(tuberculosis infection) |
| **3**  3,743 | #1 AND #2 |
| **4**  3,564 | #1 AND #2 and English |
| Total | 3564 |

**Cochrane Central Register of Controlled Trials (CENTRAL)**

| **Search Number**  Results | Search Terms |
| --- | --- |
| **1**  7814 | (mycobacterium tuberculosis):ti,ab,kw OR (tuberculosis):ti,ab,kw OR (latent tuberculosis):ti,ab,kw OR (tuberculosis infection):ti,ab,kw |
| **2**  449 | MeSH descriptor: [Mycobacterium tuberculosis] explode all trees |
| **3**  197 | MeSH descriptor: [Latent Tuberculosis] explode all trees |
| **4**  90828 | (pregnan*):ti,ab,kw OR (puerper*):ti,ab,kw OR (parturi*):ti,ab,kw OR (postpartum):ti,ab,kw OR (antepartum):ti,ab,kw OR (obstetric*):ti,ab,kw |
| **5**  7814 | #1 OR #2 OR #3 |
| **6**  358 | #4 AND #5 |
| Total | 358 |
