## Supplementary material for "*Mycobacterium tuberculosis* infection in pregnancy: a systematic review": S2 Modified NOS

**Appendix S1. Newcastle-Ottawa Scale (Modified Version)**

**Conflicts of Interest**

No evidence of conflicts of interest by one or more authors

1. Yes
0. No

**Selection**

Representative of the exposed cohort

1. Yes

0. No

Cohort characteristics are well described.

1. Yes

0. No

Ascertainment of the exposure is well described.

1. Yes

0. No

**Comparability**

If heterogeneous groups were included in the study cohort, results were disaggregated by these characteristics.

1. Yes

0. No

**Outcome**

Ascertainment of outcome is well described.

1. Yes

0. No

Was follow-up long enough for outcomes to occur?

1. Yes

0. No

Adequacy of follow-up of cohort - follow-up of ≥80% of participants

1. Yes

0. No

**Overall study quality score**

0-2 Poor

3-5 Fair

6-8 Good
