## Supplementary material for "*Mycobacterium tuberculosis* infection in pregnancy: a systematic review": S3 Characteristics of the Included Studies

| No. | Author, year | Country/ies | Years of study recruitment | Study Design | Sample size n | Included outcomes | Study quality |
| --- | --- | --- | --- | --- | --- | --- | --- |
| 1 | Arvidsson et al., 2023 | Sweden | 2013-2018 | Retrospective cohort | 439 | Cascade of care, Treatment | Good |
| 2 | Batshake et al., 2023 | Sweden | 2014-2018 | Retrospective cohort | 7,638 | Prevalence, Natural history | Good |
| 3 | Walles et al., 2023 | Sweden | 2014-2018 | Cross-sectional | 7,408 | Prevalence | Good |
| 4 | Chalid et al., 2022 | Indonesia | 2018 | Cross-sectional | 90 | Test performance | Good |
| 5 | Kaplan et al., 2022 | Kenya | 2018-2019 | Cross-sectional | 400 | Prevalence, Test performance | Good |
| 6 | Mathad et al., 2022 | Haiti, Kenya, Malawi, Thailand, and Zimbabwe | 2017-2018 | Open-label trial | 50 | Cascade of care, Treatment | Good |
| 7 | Bhosale et al., 2021 | India | 2016-2019 | Prospective cohort | 165 | Test performance, Natural history | Good |
| 8 | Bongomin et al., 2021 | Uganda | 2020 | Cross-sectional | 261 | Prevalence, Test performance | Good |
| 9 | Walles et al., 2021 | Ethiopia | 2015-2016 | Cross-sectional | 1,834 | Prevalence | Good |
| 10 | Froberg et al., 2020 | Sweden | 2016-2017 | Retrospective cohort | 4,178 | Prevalence, Cascade of care | Good |
| 11 | Jansson et al., 2020 | Sweden | 2017-2018 | Grounded theory | 16 | Cascade of care | Good |
| 12 | Jonsson et al., 2020 | Sweden | 2005-2013 | Retrospective cohort | 649,342 | Natural history | Good |
| 13 | Yassin et al., 2019 | Sudan | 2015 | Cross-sectional | 249 | Prevalence | Fair |
| 14 | Bullarbo et al., 2018 | Sweden | 2008-2012 | Retrospective cohort | 902 | Prevalence, Cascade of care | Fair |
| 15 | Moro et al., 2018 | USA, Canada | n/s | Randomised open-label trial | 125 | Treatment | Good |
| 16 | Walles et al., 2018 | Ethiopia | 2015-2016 | Cross-sectional | 829 | Prevalence, Test performance | Good |
| 17 | Vaidya et al., 2017 | India | 2011-2014 | Cross-sectional | 538 | Prevalence, Test performance | Good |
| 18 | Rendell et al., 2016 | Mongolia | 2013 | Retrospective cohort Study | 104 | Natural History | Good |
| 19 | Molina et al., 2016 | USA | 2013-2014 | Cross-sectional | 141 | Prevalence, Test performance, Cascade of care | Good |
| 20 | Gebreegziabiher et al., 2014 | Ethiopia | 2011-2012 | Cross-sectional | 85 | Prevalence, Test performance | Good |
| 21 | Mathad et al., 2014 | India | 2011-2012 | Prospective cohort | 401 | Prevalence, Test performance | Good |
| 22 | Lighter-Fisher et al., 2012 | USA | n/s | Cross-sectional | 140 | Prevalence, Test performance | Good |
| 23 | Zenner et al., 2012 | UK | 1996-2008 | Retrospective cohort | 192, 801 | Natural history | Good |
| 24 | Worjoloh et al., 2011 | USA | 2009-2010 | Cross-sectional | 220 | Prevalence, Test performance, Cascade of care | Good |
| 25 | Chehab et al., 2010 | USA | n/s | Cross-sectional | 102 | Test performance | Fair |
| 26 | Meints et al., 2010 | USA | 2003-2006 | Cross-sectional | 387 | Prevalence | Fair |
| 27 | Sheriff et al., 2010 | Tanzania | 2008 | Cross-sectional | 286 | Prevalence, Cascade of care | Good |
| 28 | Schwartz et al., 2009 | USA | 2001-2006 | Retrospective cohort | 4,049 | Prevalence, Cascade of care | Good |
| 29 | Kwara et al., 2008 | USA | 2003 | Retrospective cohort | 345 | Cascade of care | Good |
| 30 | Sackoff et al., 2006 | USA | 1999-2000 | Cross-sectional | 730 | Prevalence, Cascade of care | Good |
| 31 | Cruz et al., 2005 | USA | 2000 | Retrospective cohort | 1,331 | Prevalence, Cascade of care | Fair |
| 32 | Crampin et al., 2004 | Malawi | 1996-2001 | Case-control | 1,590 | Natural history | Good |
| 33 | Jackson et al., 2001 | USA | 2000 | Cross-sectional | 30 | Prevalence | Good |
| 34 | Medchill, 1999 | USA | 1993-1997 | Cross-sectional | 1,763 | Prevalence, Cascade of care | Good |
| 35 | Nolan et al., 1997 | USA | 1994-1995 | Cross-sectional | 1,621 | Prevalence | Poor |
| 36 | Espinal et al., 1996 | Dominican Republic | 1992-1994 | Case-control | 662 | Natural history | Fair |
| 37 | Magann et al., 1996 | USA | n/s | Cross-sectional | 1,000 | Prevalence | Good |
| 38 | Sepulveda et al., 1995 | Chile | n/s | Cross sectional | 840 | Prevalence | Fair |
| 39 | Metersky et al., 1993 | USA | 1990-1991 | Cross-sectional | 1,412 | Prevalence, Cascade of care | Good |
| 40 | Franks et al., 1989 | USA | 1981-1982 | Retrospective cohort | 3,681 | Cascade of care, Treatment | Fair |
| 41 | Plauche et al., 1983 | USA | 1981-1982 | Cross-sectional | 1,935 | Prevalence, Cascade of care | Poor |
| 42 | Present et al., 1975 | USA | n/s | Prospective cohort | 226 | Test performance | Fair |
| 43 | Bailey et al., 1972 | USA | 1970 | Cross-sectional | 522 | Prevalence, Cascade of care | Fair |
| 44 | Montgomery et al. 1968 | USA | 1960-1966 | Prospective cohort | 2,643 | Prevalence, Cascade of care | Fair |
| 45 | Edge, 1952 | USA | 1945-1946 | Retrospective cohort | 462 | Natural history | Fair |
| 46 | Seid, 1945 | USA | 1940-1942 | Cross-sectional | 1,000 | Prevalence, Cascade of care | Fair |
| 47 | Ianne et al., 1939 | USA | n/s | Retrospective cohort | 805 | Prevalence, Cascade of care | Fair |
