## Supplementary material for "*Mycobacterium tuberculosis* infection in pregnancy: a systematic review": S4 Table 1 Prevalence

|  | Author, year | Country | Years of study recruitment | Sample size | Approx. TB incidence per 100,000/annum [1,2] | Ethnicity/COB as per authors % | Age y | HIV+ n (%) | BCG Vaccinated n (%) | TST method and cut-off | TST+ (%) | IGRA Method* | IGRA+ n (%) | Total prevalence %** |
| --- | --- | --- | --- | --- | --- | --- | --- | --- | --- | --- | --- | --- | --- | --- |
| 2 | Batshake et al., 2023 | Sweden | 2014-2018 | 7,638 | 5 | Africa – 40.0, Asia – 51.0, Europe - 7.1, South America - 1.6, Other - 0.3 | Mean: 30.8 (Range 14.6-52.3) | 44 (0.6) | n/s |  |  | QuantiFERON-TB | 1,424 (18.6) | 18.6 |
| 3 | Walles et al., 2023 | Sweden | 2014-2018 | 7,408 | 5 | Africa – 45.8, Asia - 46.8, Europe – 6.0, North America - 0.1, Oceania – 0, South America - 0.9, NA - 0.1 | Mean: 29.4 | 0 | n/s |  |  | QuantiFERON-TB GOLD IN-TUBE or GOLD PLUS | 1408 (19) | 19.0 |
| 5 | Kaplan et al., 2022 | Kenya | 2018-2019 | 400 | 292 | n/s | Median: 26.0 (IQR 22.0-29.0) | 200 (50) | n/s | Mantoux,  ≥5 mm | 46 (11.6) | QFT-Plus | 66 (32.3) | 35.8 |
| 8 | Bongomin et al., 2021 | Uganda | 2020 | 261 | 199 | n/s | Median: 26.0 (IQR 23.0-30.0) | 13 (5.0) | 177 (67.8) |  |  | QuantiFERON TB Gold-Plus | 99 (37.9) | 37.9 |
| 9 | Walles et al., 2021 | Ethiopia | 2015-2018 | 1,834 | 192 | n/s | Median: 25.0 (IQR 22.0-28.0) | 170 (9.3) | n/s |  |  | QuantiFERON-TB Gold-Plus | 594 (32.4) | 32.4 |
| 10 | Froberg et al., 2020 | Sweden | 2016-2017 | 4,178 | 6 | n/s | n/s | 39 (0.9) | n/s |  |  | n/s | 862 (20.6) | 20.6 |
| 13 | Yassin et al., 2019 | Sudan | 2015 | 249 | 88 | n/s | Mean: 29.6 | n/s | n/s |  |  | QuantiFERON-TB Gold In-Tube | 45 (18.1) | 18.1 |
| 14 | Bullarbo et al., 2018 | Sweden | 2008-2012 | 902 | 6 | n/s | Range: 10.0-45.0 | n/s | n/s | Mantoux,  ≥10 mm | 327 (36.0) |  |  | 36.0 |
| 16 | Walles et al., 2018 | Ethiopia | 2015-2016 | 829 | 177 | n/s | n/s | 49 (5.9) | n/s |  |  | QuantiFERON-TB Gold-Plus | 277 (33.0) | 33.0 |
| 17 | Vaidya et al., 2017 | India | 2011-2014 | 538 | 268 | n/s | n/s | 243 (45) | n/s |  |  | QuantiFERON-TB GOLD IN-TUBE | 162 (30.0) | 30.0 |
| 19 | Molina et al., 2016 | USA | 2013-2014 | 141 | 3 | Hispanic/Latina – 50.0; African/African American - 27.0; Asian – 13.0; White - 9.0; Other – 1.0 | Median: 28.0 (IQR 24.6-33.1) | 4 (2.8) | 74 (52.0) | Mantoux,  n/s | 6 (4.2) | T-SPOT.TB | 3 (2.2) | 5.0 |
| 20 | Gebreegziabiher et al., 2014 | Ethiopia | 2011-2012 | 85 | 239 | n/s | Median: 25.0 (IQR 22.0-28.0) | n/s | 13 (15.3) |  |  | QuantiFERON-TB GOLD IN-TUBE | n/s (26.8) | 26.8 |
| 21 | Mathad et al., 2014 | India | 2011-2012 | 401 | 268 | n/s | n/s | 0 |  | Mantoux,  ≥10 mm | 59 (14.0) | QuantiFERON-TB GOLD IN-TUBE | 150 (37.0) | 37.0 |
| 22 | Lighter-Fisher et al., 2012 | USA | n/s | 140 |  | Born in TB endemic region – 41.0 | Mean: 18.5 (Range 13.5-36.5) | 0 | 56 (40.0) | Mantoux,  ≥10 mm | 28 (20.0) | QuantiFERON-TB GOLD IN-TUBE | 15 (10.7) | 20.0 |
| 24 | Worjoloh et al., 2011 | USA | 2009-2010 | 220 | 4 | White - 8.5; Hispanic 69.8; American Indian 1.0; Asian - 8.0 | Mean: 25.0 (Range 17.0-41.0) | 0 |  | Mantoux,  ≥10 mm | 45 (23.0) | QuantiFERON-TB GOLD IN-TUBE | 28 (14.0) | 23.0 |
| 26 | Meints et al., 2010 | USA | 2003-2006 | 387 | 5 | African – 29.0; European – 23.0; Latin American – 22.0; Asian – 21.0; Middle Eastern – 14.0 | n/s | n/s | n/s | n/s | 82 (21.2) |  |  | 21.2 |
| 27 | Sheriff et al., 2010 | Tanzania | 2008 | 286 | 474 | n/s | Mean: 25.0 (Range 16.0-41.0) | 13 (4.5) | n/s | Mantoux,  ≥5 mm | 87 (30.4) |  |  | 30.4 |
| 28 | Schwartz et al., 2009 | USA | 2001-2006 | 4049 | 5 | Asian - 49.6; Caucasian - 1.4; Hispanic - 42.9 | Mean: 27.0 (Range 13.0-46.0) | 0 | n/s | n/s,  ≥10 mm | 1,935 (50.4) |  |  | 50.4 |
| 30 | Sackoff et al., 2006 | USA | 1999-2000 | 678 | 7 | Caribbean – 36.0; Central America – 31.0; Asia- 18.0; South America – 13.0 | Median: 26.0 (IQR 22.0-31.0) | 0 | n/s | n/s,  ≥10 mm | 341 (50.3) |  |  | 50.3 |
| 31 | Cruz et al., 2005 | USA | 2000 | 1,331 | 7 | n/s | n/s | n/s | n/s | n/s | 425 (31.9) |  |  | 31.9 |
| 33 | Jackson et al., 2001 | USA | 2000 | 30 | 7 | White – 47.0 | Mean: 27.5 | 0 | n/s | Mantoux,  ≥10 mm | 0.0 |  |  | 0.0 |
| 34 | Medchill, M.T., 1999 | USA | 1993-1997 | 1,763 | ̴45 (New York) | White – 38.0; Hispanic – 54.0; Black 4.0; Asian 1.0; Native America- 1.4; Other - 1.6 | n/s | n/s | n/s | Mantoux,  ≥15 mm | 246 (15.2) |  |  | 15.2 |
| 35 | Nolan et al., 1997 | USA | 1994-1995 | 1,621 | ̴32 (New York) | African American - 78.6; White - 9.7; Hispanic - 5.6; Other - 5.9 | n/s | n/s | n/s | Mantoux,  ≥10 mm | 71 (4.4) |  |  | 4.4 |
| 37 | Magann et al., 1996 | USA | n/s | 1,000 |  | Caucasian - 58.5; African American – 12.0; Asian - 18.7; Hispanic - 10.8 | Mean: 25.3 (Range 13.0-42.0) | n/s | 0 | Mantoux,  ≥5 mm | 43 (4.3) |  |  | 4.3 |
| 38 | Sepulveda et al., 1995 | Chile | n/s | 840 |  | n/s | n/s | 0 | 807 (96.0) | n/s,  ≥10 mm | (51.0– 57.0) |  |  | 51.0– 57.0 |
| 39 | Metersky et al., 1993 | USA | 1990-1991 | 1,412 | ̴48 (New York) | n/s | n/s | n/s | n/s | Mantoux,  ≥5 mm | 259 (18.3) |  |  | 18.3 |
| 41 | Plauche et al., 1983 | USA | 1981-1982 | 1,935 | ̴22 (New York) | n/s | n/s | n/s | n/s | Mantoux,  ≥10 mm | 85 (4.4) |  |  | 4.4 |
| 42 | Bailey et al., 1972 | USA | 1970 | 522 | ̴33 (New York) | n/s | Range 15.0-40.0 | n/s | n/s | n/s,  ≥5 mm | 25 (4.8) |  |  | 4.8 |
| 45 | Montgomery et al. 1968 | USA | 1960-1966 | 2,357 | ̴60 (New York) | African American – 100.0 | n/s | n/s | n/s | n/s,  ≥10 mm | 199 (9.6) |  |  | 9.6 |
| 46 | Seid, M.J., 1945 | USA | 1940-1942 | 1,000 | ̴120 (New York) | n/s | Range 16.0-43.0 | n/s | n/s | n/s | 380 (38.0) |  |  | 38.0 |
| 47 | Ianne et al., 1939 | USA | n/s | 805 |  | n/s | n/s | n/s | n/s | n/s | 284 (41.1) |  |  | 41.1 |
|  | Abbreviations: IQR = Interquartile range; HIV+ = Human Immunodeficiency Virus seropositive; BCG= Bacille Calmette-Guérin; TST = Tuberculin skin test; IGRA = Interferon-Gamma Release Assay; n/s= not stated  *Manufacturer cut-offs were used for all studies **Positive by either IGRA or TST | | | | | | | | | | | | | |
